# Impact of climatic variations on the global distribution and prevalence of Mpox disease: Analysis of data from 52 countries using linear regression and time forecasting analyses

**DOI:** 10.1101/2025.02.04.25321656

**Authors:** Iyanuloluwa S. Ojo, Olurotimi John Badero, Muhammad Aziz Rahman, Nicholas Aderinto, Onur Oral, Rishad Choudhury Robin, Emmanuel Jesuyon Dansu, Temitope Ogunjimi, Faridat Ibidun

**Author notes:** LinkedIn: https://www.linkedin.com/in/iyanuloluwa-ojo-b2b1b1137/.

## Abstract

MPOX infection has become a global health concern. Despite this global concern, the disease has shown epidemiological variations across different countries. This study examines the impact of climatic variations on MPOX prevalence and mortality using quantitative, cross-sectional approach. We analyzed daily confirmed MPOX cases from 52 countries using linear regression and Prophet models, focusing on average mean surface air temperature, precipitation, and climate types based on the Köppen classification. Globally, no significant relationship was found between temperature, precipitation, and MPOX prevalence (p-values 0.99 and 0.82); no significance was also noted for mortality globally (p-values 0.45 and 0.54). However, temperature had a more substantial impact on MPOX prevalence in Europe and South America (p-values of 0.049 and 0.0036, respectively). Precipitation was also significant in South America (p-value 0.044). The Prophet model, accounting for seasonality, revealed fluctuating influences of temperature and precipitation on MPOX prevalence, with their effects diminishing over time. Tropical and temperate climate types were most strongly associated with increased MPOX transmission. Monitoring these climatic factors, particularly in affected regions, is crucial for understanding and mitigating MPOX spread and its associated mortality.

## 1. INTRODUCTION

Mpox was declared a public health emergency of international concern by WHO Director-General Dr. Tedros Adhanom Ghebreyesus in August 2024 [1]. This outbreak is part of recurring cycles, with the first multi-country outbreak starting in 2022 and being declared an emergency [1], [2]. Initially isolated from captive monkeys in 1959, the virus was first identified in humans in the Democratic Republic of Congo (DRC), which is the epicenter of the 2024 outbreak [3], [4]. As of May 1, 2024, Mpox has caused 225 deaths out of 103,429 cases globally [5], though this figure may be an underestimation, with reports suggesting over 1,000 deaths in the DRC alone [6], [7]. In non-endemic regions, where the virus is new, the population lacks pre-existing immunity, which can lead to more widespread transmission and potentially more severe outcomes [8]. While monkeypox is often self-limiting in healthy individuals, immunocompromised patients may experience more severe or prolonged illness [9]. There are two major strains: Clade 1, responsible for the 2024 outbreak in Africa, and Clade 2, which caused the 2022 outbreak in Europe [10], [11]. Mpox primarily spreads through close contact with infected animals or humans, especially through sexual contact [12], [13], [15], [16]. Symptoms include a rash with blisters or ulcers, fever, backaches, joint pain, exhaustion, and swollen lymph nodes, while some patients have been noted to have anal ulcerations [14], [15], [16], [17]. Viral infections and contact diseases have been shown to be influenced by climatic variations. Seasonal oscillations in a pathogen’s effective reproductive number have been observed for some viral infections, possibly driven by wintertime survival or relative wintertime immunosuppression [18]. For example, the influenza virus is more active in low daily temperatures, with a high incidence during cooler temperatures in temperate regions [19], [20]. Similarly, COVID-19, caused by the SARS-CoV-2 virus, was found to be modestly curbed by high temperatures [21]. Other studies suggested that COVID-19 transmission could be higher at an absolute humidity of 5-10g/m³ [22], [23]. These findings suggest that climatic factors might influence the increasing prevalence of certain viral contact diseases.

The diverse and ambiguous distribution of MPOX raises questions about the factors influencing this pattern. For instance, the differing prevalence of MPOX between Kenya and Trinidad and Tobago is noteworthy. Kenya recorded 12 confirmed cases of MPOX, compared to Trinidad and Tobago’s 106 confirmed cases in 2024, despite Kenya’s proximity to the outbreak’s epicentre [7]. Furthermore, Trinidad and Tobago has a higher GDP per capita and adult literacy rate than Kenya, making the prevalence findings puzzling [24], [25]. Another curious observation is that no MPOX cases have been reported in countries like Morocco, Egypt, and Libya, while countries like Spain, Brazil, and China have seen a sharp rise in confirmed cases, despite the former being geographically closer to the outbreak’s epicentre [25]. There have been several instances of neighbouring countries with significantly different confirmed cases of MPOX. For example, Spain has recorded over 300 confirmed cases of MPOX while Morocco has none. Moreover, despite the Democratic Republic of Congo being the source of the outbreak, its eastern neighbour Rwanda has only recorded a few MPOX cases. While other factors might be involved in this differing prevalence between these countries, the fact that these countries have relatively different climate types suggests that climatic variations could play a role in explaining these differences and thus warrants further study.

The potential influence of climatic factors on the prevalence and transmission of MPOX disease has been understudied. The combination of climate, environmental conditions, and human interactions with their surroundings can create conditions that increase the spread and impact of MPOX [26]. Since the 2022 outbreak, only one study has examined the impact of meteorological factors on MPOX prevalence, covering data from May 6, 2020, to October 10, 2022 [27]. There is a need to further investigate the influence of climatic variations on MPOX prevalence, particularly in the context of the 2024 outbreak, given the disease’s diverse and ambiguous global distribution.

The aim of the study is to determine the influence of climatic variations, particularly the average mean surface air temperature and precipitation, on the prevalence of MPOX and the mortality associated with it.

## 2. MATERIALS AND METHODS

### 2.1 Daily MPOX Confirmed Cases and Mortality in 2024

We extracted 2024 MPOX confirmed cases and mortality data primarily from *Our World in Data*, the regional WHO bulletin for Africa, and the Joint ECDC-WHO Regional Office for Europe report [5], [28], [29]. This data includes the daily recorded number of patients confirmed with MPOX via a polymerase chain reaction (PCR) test and the number of deaths from MPOX per day.

Additionally, we obtained data from several reputable news agencies across diverse countries when information was not available from the primary sources. The data were collected from January 1, 2024, to August 23, 2024.

### 2.2 Climatic Factors

We utilized two important climatic factors: the average mean surface air temperature and precipitation in the form of rain, sleet, and snow. Data for each assessed country was extracted from the World Bank Climate Change Knowledge Portal [30]. This data was based on the climate context for the current climatology, 1991-2020, derived from observed historical data.

We also used the Köppen Climate Classification types, derived from the National Geographic Society [31].

### 2.3 Other Factors Assessed

In order to limit omitted variable bias, we decided to also assess the impact of some factors outside the scope of our study.

We derived the Adult Literacy Data from the UNESCO Institute for Statistics via the World Bank (2023) and Our World in Data [25], [32]. UNESCO defines literacy as the ability to read and write basic everyday statements. However, literacy standards differ across countries, and data from North America and Western Europe, which rely on more thorough assessments, cannot be easily compared with global data and could not be obtained.

Net Migration Rate data was derived from the CIA’s *The World Factbook*, and the Gross Domestic Product (GDP) per capita was obtained from the International Monetary Fund [24], [33].

### 2.4 Linear Regression Model

A linear regression model was used in R to determine whether there is a linear relationship between the climatic factors extracted for each country and the prevalence and mortality of MPOX, both globally and continentally. The R package ‘ggplot2’ and the function ‘lm’ were employed to analyze the data using the linear regression model defined as:

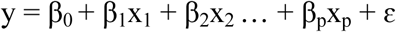

Where y represents MPOX prevalence or mortality, x_1_ and x_2_…x_p_ are the predictors (AMSAT and precipitation), β_0_ is the intercept of the regression line, β1, β_2_…β_P_ represent the strength and direction of the effect of each predictor on y, while ɛ accounts for variability not captured by the predictors.

### 2.5 Time Forecasting Model

The automatic forecasting time series model, Prophet, was used in R to analyze the impact of climatic variations on MPOX prevalence globally. Developed by Meta, Prophet is primarily designed for time forecasting but can also be adapted to assess the influence of one variable on another. We chose Prophet for its robustness in handling time series data with seasonality and irregular patterns. Average mean surface air temperature and precipitation were added as external regressors to examine their impact on MPOX prevalence.

The Prophet model was trained using each external regressor, generating forecasts for MPOX prevalence while accounting for seasonality. The R package ‘future’ was utilized to parallelize computations and ensure optimal efficiency during model training, while ‘ggplot2’ was employed to visualize the results. This included the forecast plot, trend and seasonal plots, and the regressor plot, which is of primary concern in our study. The analysis is expressed as:

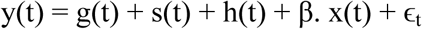

Where y(t) is the predicted value at time t, g(t) is the trend function, s(t) is the seasonal function, h(t) is the effect of other events, β is the coefficient showing the impact of the regressor, and x(t) is the value of the regressor at time t.

### 2.6 Inclusion Criteria

For both the linear regression and time forecasting analyses, it is essential that a reliable dataset is available for the countries included in the study.

For the time-forecasting analysis, selected countries must have reported at least 10 confirmed MPOX cases and must not be neighbouring the Democratic Republic of Congo, the epicentre of the 2024 outbreak and the most affected country.

These inclusion criteria were established to minimize selection bias as effectively as possible.

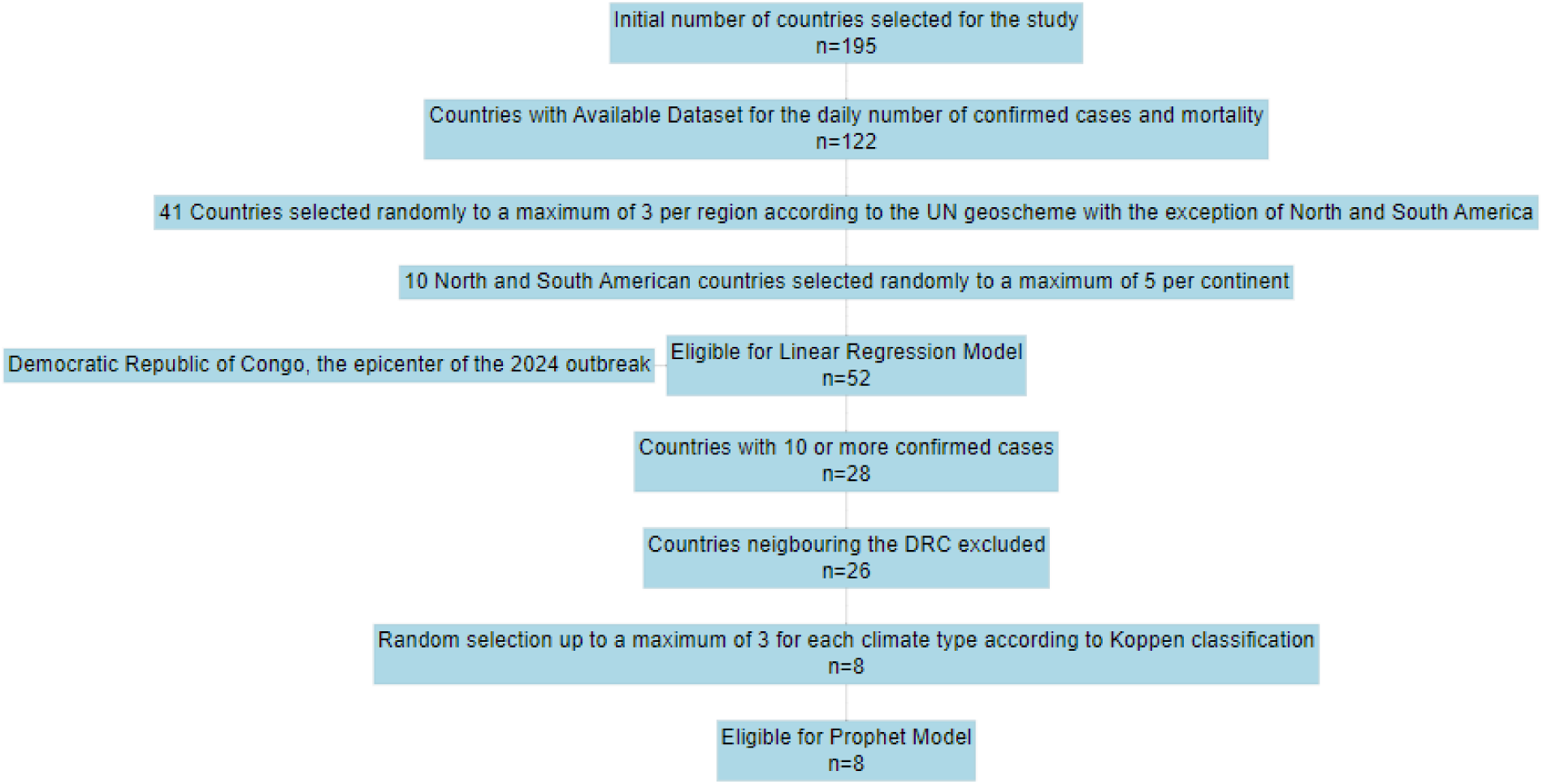
PRISMA-STYLED DIAGRAM SHOWING THE SELECTION PROCESS BASED ON THE INCLUSION CRITERIA.

### 2.7 Statistical Analysis

We collated data using WPS Office Excel version 2021 and analyzed it using R software version 4.4.1. A linear regression model was employed to determine the relationship between climatic factors and MPOX prevalence and mortality in 2024. Additionally, the model assessed the impact of other factors such as the adult literacy rate, GDP per capita, and net migration rate.

The evolution of MPOX infection was examined using the Automatic Forecasting Time-Series Model, Prophet, which accounts for seasonality—an aspect not considered by the linear regression model.

In all models, climatic factors served as the dependent variables, while MPOX prevalence was the independent variable. The number of deaths from MPOX was also considered globally as an independent variable in the linear regression model. The relationship between these variables was verified using coefficients, the 95% confidence interval, the F-statistic, and the corresponding p-value.

## 3. RESULTS

Table 3.1 shows the descriptive statistics of both the independent and dependent variables. The average mean surface air temperature globally was 18.03-degrees Celsius with a standard deviation of 8.25, a minimum of −4.03 which was recorded for Canada and a maximum of 28.17 which was recorded for the United Arab Emirates. The mean precipitation was 1049.06 with a standard deviation of 734.21, a minimum of 19.62 recorded for Egypt, and a maximum of 2996.28 recorded for Malaysia. An average of 140.42 cases were confirmed to be MPOX via the polymerase chain reaction (PCR) with a standard deviation of 495.74. The lowest number of cases confirmed was zero, which was recorded for multiple countries, while the highest number was 3235 for the Democratic Republic of the Congo, the source of the 2024 outbreak. The average number of mortality cases was 10.039 with a standard deviation of 70.84; the minimum number of deaths was zero, which was recorded for 88.5% of the countries studied, while the maximum was 511, which was recorded for the Democratic Republic of the Congo.

**Table 3.1:**
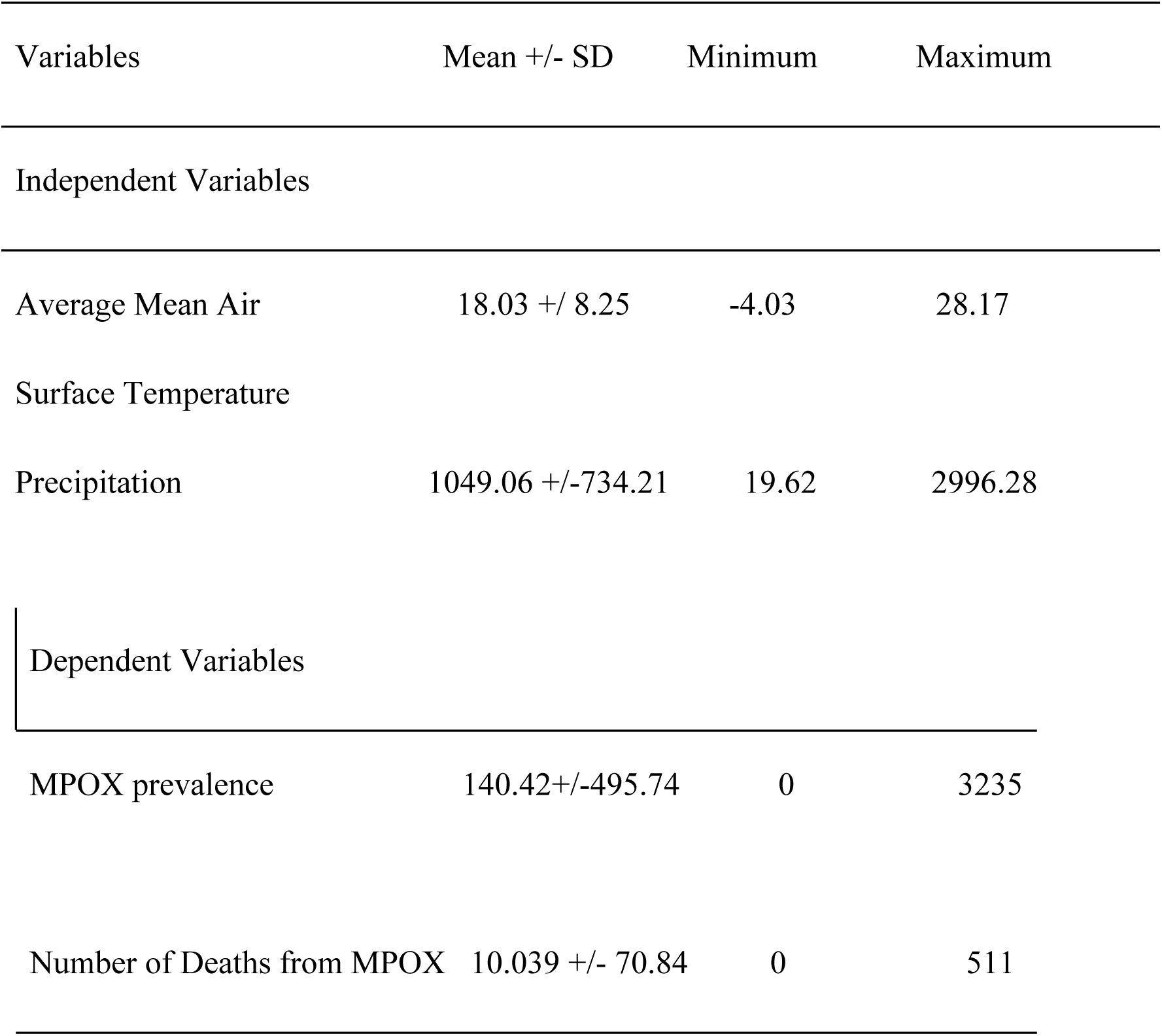
Descriptive statistics for the climatic factors, MPOX prevalence and mortality from MPOX.

| Variables | Mean +/- SD | Minimum | Maximum |
| --- | --- | --- | --- |
| Independent Variables |  |  |  |
| Average Mean Air | 18.03 +/- 8.25 | -4.03 | 28.17 |
| Surface Temperature |  |  |  |
| Precipitation | 1049.06 +/-734.21 | 19.62 | 2996.28 |
| Dependent Variables |  |  |  |
| MPOX prevalence | 140.42+/-495.74 | 0 | 3235 |
| Number of Deaths from MPOX | 10.039 +/- 70.84 | 0 | 511 |

The linear regression model analysis in Table 3.2 shows that the average mean surface air temperature and precipitation have no significant effect on the MPOX prevalence **globally,** with a 95% confidence interval of −17.17 to 16.98 and −0.17 to 0.21 respectively, a low adjusted R-squared of −0.020 and −0.019, respectively, and a p-value >0.05 (0.99 and 0.82, respectively). The global findings are echoed in **Africa** with a 95% confidence interval of −11.45 to 5.59 and −0.035 to 0.055, respectively, a low adjusted R-squared of −0.037 and −0.069, respectively, and a p-value of 0.47 and 0.64, respectively. The global findings are also echoed in **Asia,** with a 95% confidence interval of −17.15 to 0.81 and −0.079 to 0.052, respectively, a low adjusted R-squared of 0.18 and −0.069, respectively, and a p-value of 0.071 and 0.66, respectively. However, the average mean surface air temperature has a weak impact on MPOX prevalence when compared to the global findings. **Europe** findings are slightly opposite to the global findings. The AMSAT has a positive correlation with the MPOX prevalence in Europe with a 95% confidence interval of 0.10 to 31.45, a positive adjusted R squared value, and a p-value <0.05 (0.049). Precipitation findings in Europe, however, echoed global findings with a 95% confidence interval of −0.35 to 0.20, a negative adjusted R squared value of −0.058, and a p-value >0.05 (0.54). **North America** also echoed the global findings with a 95% confidence interval of −107.55 to 70.15 and −2.19 to 1.065, respectively, a low adjusted R-squared of −0.16 and 0.050, respectively, and a p-value of 0.55 and 0.35, respectively. **In South America**, findings were of significant opposite to the global findings. Both AMSAT and precipitation have a significant positive correlation with MPOX prevalence with a 95% confidence interval of 4.43 to 9.90 and 2.765e-03 to 0.11, respectively, a positive adjusted R-squared of 0.95 and 0.72, respectively, and a p-value of 0.0036 and 0.044, respectively. Only two countries, Australia and New Zealand, met the inclusion criteria in **Oceania,** which is not enough data for a linear regression model.

**Table 3.2:** Climatic Variations associated with MPOX Prevalence using the Linear Regression Model.

| Climatic Factors | Coefficient | 95% CI | Adjusted R<sup>2</sup> | p-value |
| --- | --- | --- | --- | --- |
| GLOBALLY |  |  |  |  |
| Average Mean | 142.09 | -17.17 to 16.98 | -0.020 | 0.99 |
| Surface Air<br>Temperature |  |  |  |  |
| Precipitation | 117.22 | -0.17 to 0.21 | -0.019 | 0.82 |
| AFRICA |  |  |  |  |
| Average Mean | 92.84 | -11.45 to 5.59 | -0.037 | 0.47 |
| Surface Air<br>Temperature |  |  |  |  |
| Precipitation | 13.36 | -0.035 to 0.055 | -0.069 | 0.64 |
| ASIA |  |  |  |  |
| Average Mean | 211.19 | -17.15 to 0.81 | 0.18 | 0.071 |
| Surface Air<br>Temperature |  |  |  |  |
| Precipitation | 55.14 | -0.079 to 0.052 | -0.065 | 0.66 |
| EUROPE |  |  |  |  |
| Average Mean | -68.98 | 0.10 to 31.45 | 0.27 | 0.049 |
| Surface Air<br>Temperature |  |  |  |  |
| Precipitation | 136.22 | -0.35 to 0.20 | -0.058 | 0.54 |
| NORTH AMERICA |  |  |  |  |
| Average Mean | 656.47 | -107.55 to 70.15 | -0.16 | 0.55 |
| Surface Air |  |  |  |  |
| Temperature |  |  |  |  |
| Precipitation | 1091.44 | -2.19 to 1.065 | 0.050 | 0.35 |
| SOUTH AMERICA |  |  |  |  |
| Average Mean | -72.45 | 4.43 to 9.90 | 0.95 | 0.0036 |
| Surface Air |  |  |  |  |
| Temperature |  |  |  |  |
| Precipitation | -23.70 | 2.765e-03 to 0.11 | 0.72 | 0.044 |
| OCEANIA |  |  |  |  |
| Not enough data to analyze as only two countries met the inclusion criteria. |  |  |  |  |

**Table 3.3:**
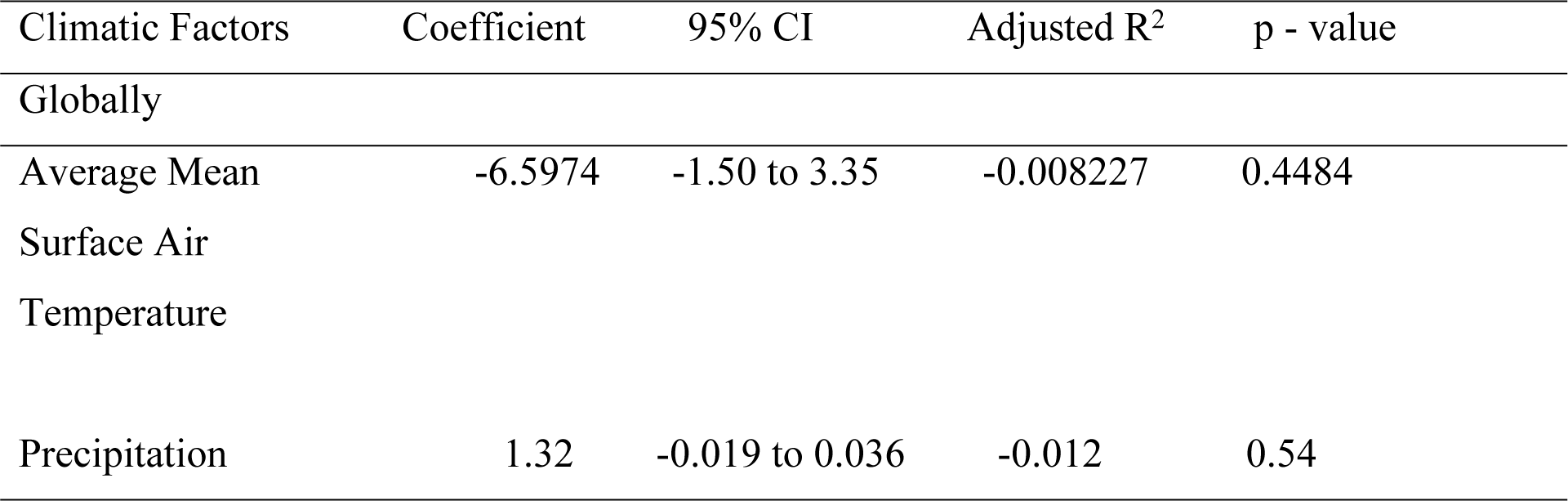
Climatic Variations associated with Mortality from MPOX using the Linear Regression Model.

**Table 3.4:** The Effect of Other Selected Factors on MPOX Prevalence.

| Other Factors | Coefficient | 95% CI | Adjusted R<sup>2</sup> | p-value |
| --- | --- | --- | --- | --- |
| Migration | 135.54 | -37.93 to 58.42 | -0.016 | 0.67 |
| Adult Literacy<br>Rate | 165.21 | -12.71 to 13.14 | -0.020 | 0.95 |
| GDP per capita | 150.32 | -4.58 to 3.97 | -0.020 | 0.89 |

The linear regression model analysis in Table 3 shows that there is no significant correlation between the climatic variations assessed and the mortality from MPOX with a 95% confidence interval of −1.50 to 3.35 and −0.019 to 0.036, negative adjusted R-squared value of −0.008227 and −0.012 and p-value of 0.4484 and 0.54 for average mean surface air temperature and precipitation, respectively.

The three other factors assessed do not have a significant influence on MPOX prevalence with a 95% confidence interval of −37.93 to 58.42, −12.71 to 13.14 and −4.58 to 3.97, negative adjusted R-squared values of −0.016, −0.020 and −0.020 and p-values of >0.05 (0.67, 0.95, 0.89) for migration, adult literacy rate and GDP per capita respectively. Based on the p-values, none of the three other factors assessed had a significant relationship with MPOX prevalence.

From L-R: CD - Continental KC D, TA - Tropical KC A, DB - Dry KC B, ED - Extremely Diverse, HH - Highland KC H, MCD - Majorly Continental KC D, MTC - Majorly Temperate KC C, MTA - Majorly Tropical KC A, PE - Polar KC E, TC-Temperate KC C Figure 3.1 shows the Linear Regression analysis comparing the impact of climate type on MPOX prevalence globally. Tropical climate type (Koppen Classification Type A) had the highest impact on MPOX prevalence, with temperate climate type (Koppen Classification Type B) and extremely diverse climate coming close. Continental climate type (Koppen Classification Type D) and Polar climate (Koppen Classification Type E) have the lowest impact. However, the p-value is 0.98, which means that climate type generally has little significance in determining MPOX prevalence globally.

**FIGURE 3.1:**
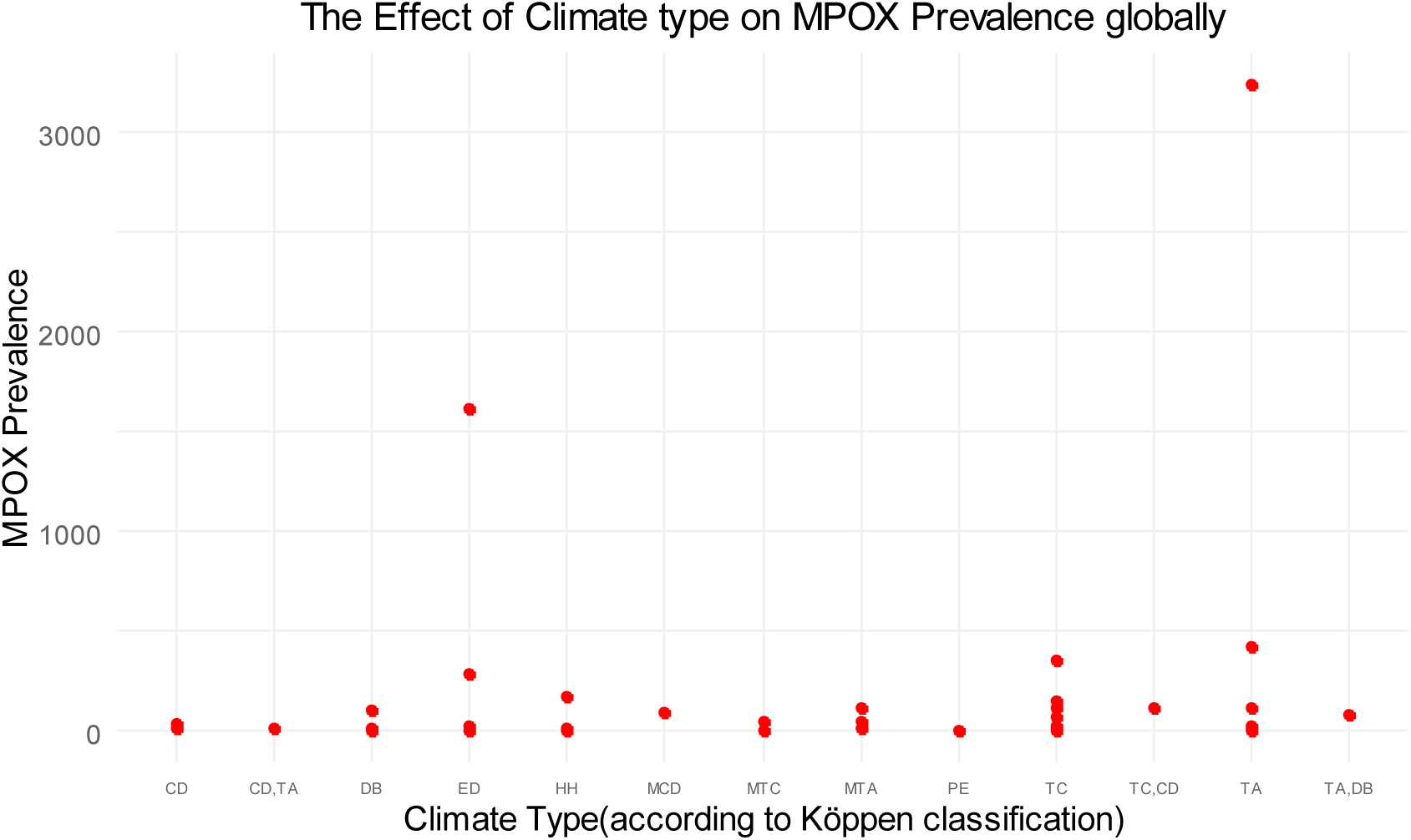
THE GLOBAL EFFECT OF CLIMATE TYPE ON MPOX PREVALENCE IN THE LINEAR REGRESSION MODEL.

Figure 3.2 shows the effect of the average mean surface air temperature on MPOX prevalence in the prophet model. The historical data up to September 2024 shows a tall, recurring pattern up to the positive threshold, which indicates that AMSAT has a strong positive correlation with MPOX prevalence at certain intervals during the observed months (January-August 2024). However, the horizontal line post historical data after October 2024 signifies a diminishing effect of AMSAT on MPOX prevalence over time.

**FIGURE 3.2:**
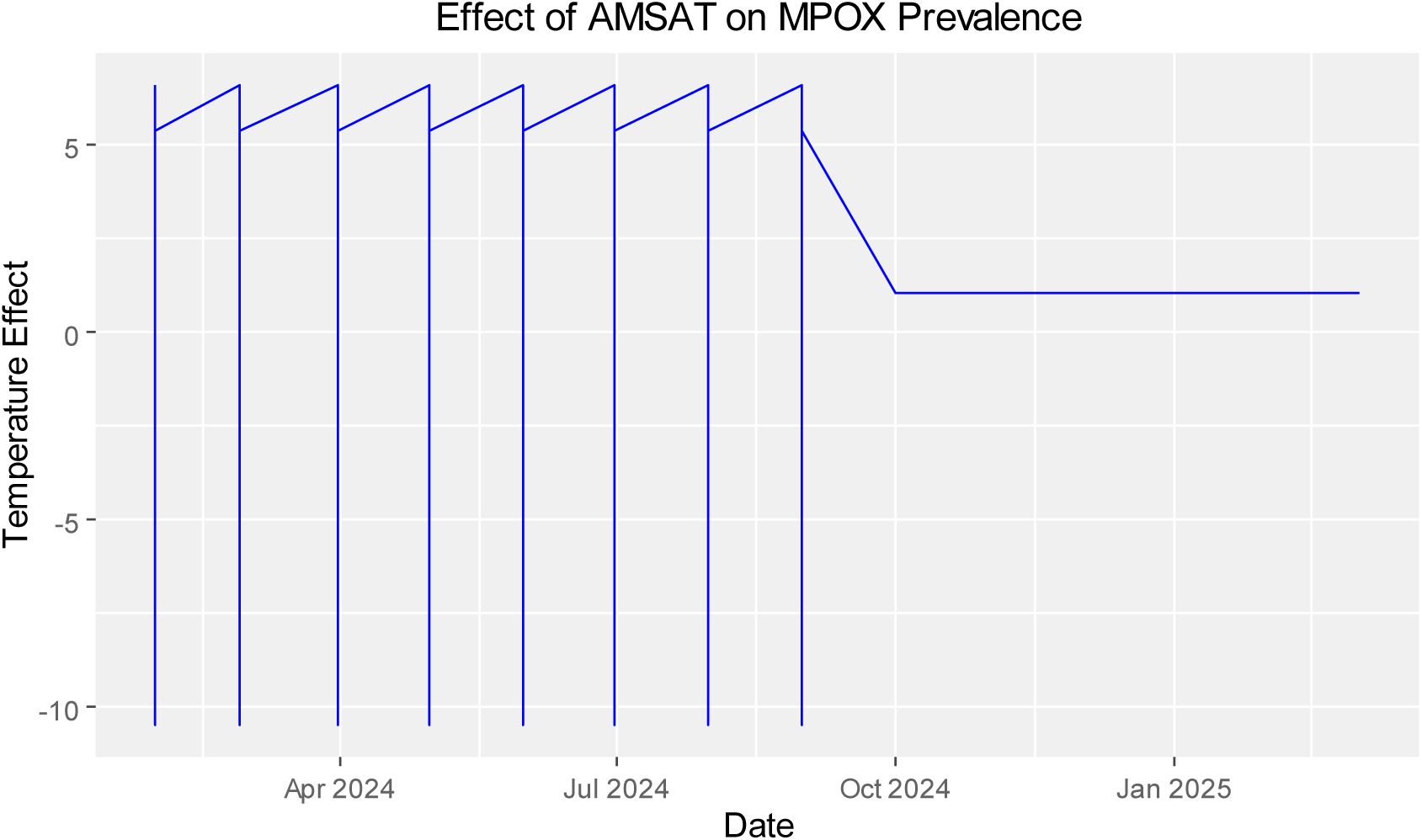
THE GLOBAL EFFECT OF THE AVERAGE MEAN SURFACE AIR TEMPERATURE ON MPOX PREVALENCE IN THE PROPHET MODEL.

Figure 3.3 shows the effect of precipitation on MPOX prevalence in the prophet model. The tall, recurring spikes span across both positive and negative fields, indicating that precipitation has a strong influence on MPOX prevalence, alternating between increasing and decreasing MPOX prevalence at certain intervals from January to September 2024. However, the horizontal line post historical data from October 2024 shows a weakening influence on MPOX prevalence over time. Given more credence to this is that the horizontal line is at the negative zone.

**FIGURE 3.3:**
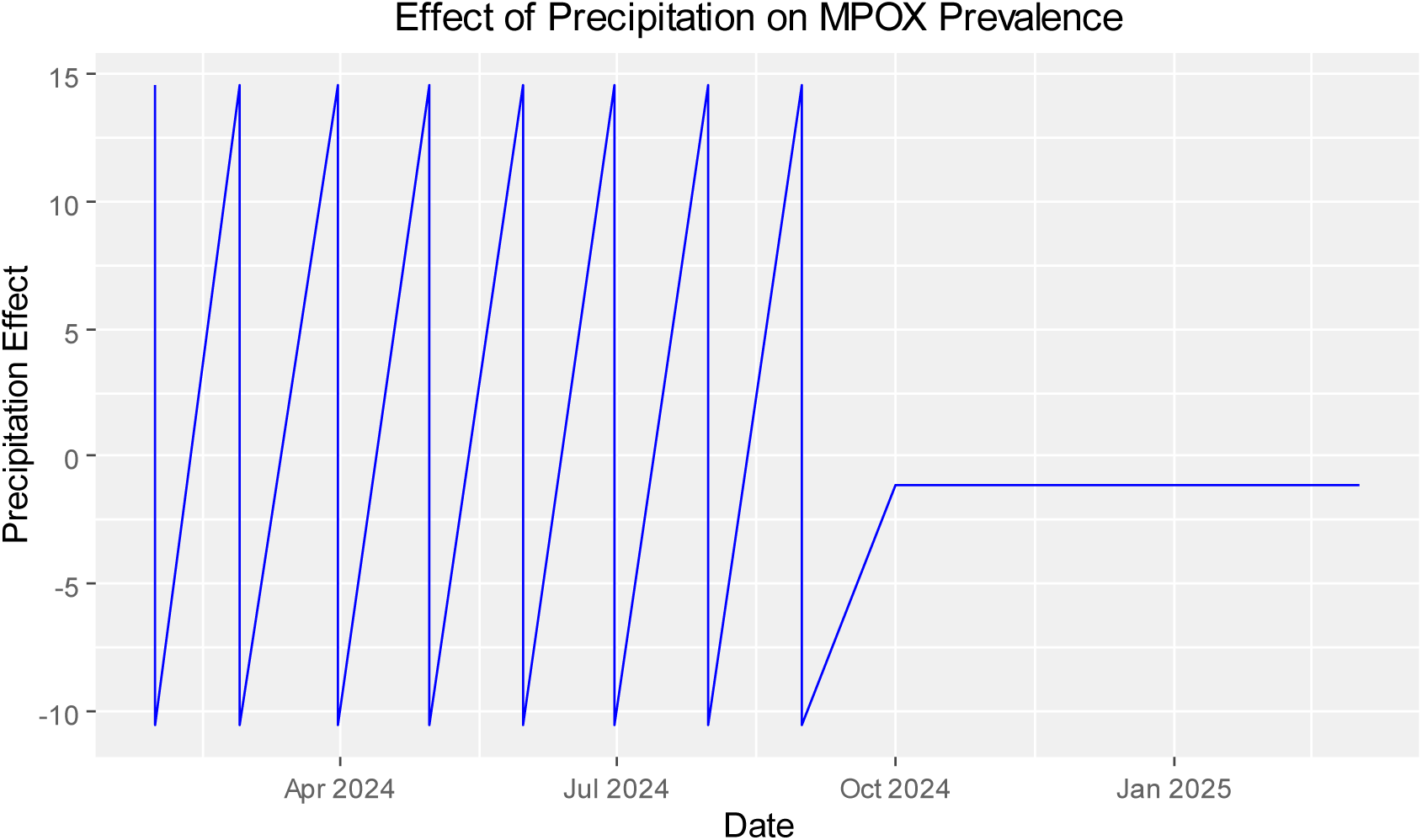
THE EFFECT OF PRECIPITATION ON MPOX PREVALENCE IN THE PROPHET MODEL.

## 4. DISCUSSION

To the best of our knowledge, this is the second study to assess the impact of climatic variations on the disruptive MPOX outbreaks and the first to do so in the context of the recent 2024 outbreak.

Both the average mean surface air temperature (AMSAT) and precipitation had no significant relationship with MPOX prevalence and mortality globally based on the linear regression model. However, AMSAT was influential on MPOX prevalence in South America and Europe, while precipitation was significant on MPOX prevalence in South America.

This could be explained by the climatic diversity on each continent. When compared to the other continents, excluding Antartica and Oceania, South America and Europe are the least diverse in climatic types. Most of South America is of the tropical and subtropical climates, which have been found to be the most favorable climate type for MPOX prevalence in this study, with the Amazon basin encompassing most of the continent [34]. Europe majorly has the temperate climate type and the continental climate types in a few countries [34]. The temperate climate type is also favorable to MPOX prevalence based on this study.

Precipitation in South America is very high, especially around the Amazon Basin. This high precipitation is a conducive environment for known vectors of the MPOX disease, such as monkeys and squirrels and can ecologically influence the MPOX disease prevalence [35].

Since climate types are extremely diverse in Africa, Asia, and North America, the varying temperature and precipitation might mask the influence of the average mean surface air temperature and precipitation on MPOX prevalence in these regions.

The Average Mean Surface Air Temperature (AMSAT) shows a positive correlation with MPOX disease in countries already affected, as indicated by the Prophet model. However, its impact on MPOX prevalence is globally weak, with a p-value of 0.99 in the linear regression model. The flat line after the historical data in the Prophet model suggests a weak relationship after September. This finding is consistent with a study by Islam et al. (2022), which also found a positive correlation between temperature and daily monkeypox cases [27]. This indicates that AMSAT might play a significant role in affecting the virulence and severity of MPOX, but it does not appear to have a notable impact on the overall incidence or occurrence of the disease. Temperature has been shown to influence the virulence of the pox virus in experiments with chick embryos [36]. However, the impact of temperature on virus virulence can vary; for example, a study on COVID-19 found that both extremely high and low temperatures significantly affected SARS-CoV-2 virulence [21]. In contrast, our study found that extreme temperatures tend to reduce MPOX prevalence, as shown in the tables. Thus, a positive correlation with temperature appears to hold true only within a specific range.

Precipitation exerts a dynamic influence in countries already affected by the MPOX virus, according to the Prophet model. However, its global influence on MPOX is weak, with a p-value of 0.82 in the linear regression model. The horizontal flat line post-historical data suggests that the impact of precipitation on MPOX prevalence might depend on other factors not accounted for in the forecasted period. This suggests that precipitation does not influence MPOX prevalence globally, which contrasts with a study showing increased diarrhoea prevalence in sub-Saharan children with low rainfall during the dry season [37], [38]. Precipitation has also been reported as a negative driver of cough, diarrhoea, and fever in another study, indicating a potential dynamic two-legged influence of precipitation, as observed in this study [39]. High precipitation beyond 1500 mm has, however, been associated with a high prevalence of MPOX in South America.

The strong effect of AMSAT on MPOX prevalence in tropical and temperate climates could also be explained by the virus’s mode of transmission. MPOX is primarily transmitted through contact with animals such as pouched rats, rope and sun squirrels, African dormice, monkeys, pigs, hedgehogs, and rodents, and secondarily through human-to-human contact, with sexual contact being prevalent among men who have sex with men [40]. It can also be transmitted via contaminated surfaces or cloth. The significant impact of temperature on MPOX prevalence in already affected countries might be due to increased sympathetic activity, which activates cholinergic fibres supplying sweat glands, leading to increased sweating and a higher chance of contact with contaminated items [41]. MPOX potential for sustained human-to-human transmission through close contact and contaminated materials raises significant concerns and should not be underestimated [42]. High temperatures have also been reported to enlarge skin pores [43]. Conversely, precipitation does not affect MPOX transmission as AMSAT does. However, extremely high precipitation (rainfall) is conducive for vectors of the MPOX disease, particularly monkeys, while low precipitation can cause dehydration and dry skin, making it harder for the virus to penetrate the skin barrier. This explains why high precipitation in South America strongly influences MPOX prevalence.

Our findings have significant implications. They elucidate the varying prevalence of MPOX across countries with similar net migration rates, economic influences, and literacy rates. For example, despite being close geographically, Spain and Morocco show different MPOX cases. Spain, with a temperate climate and a precipitation level of 623.07 mm, has recorded 352 MPOX cases in 2024. In contrast, Morocco, with a dry climate and a lower precipitation level of 315.87 mm, has not reported any MPOX cases in 2024. Libya, a neighbouring country to Morocco with similar climatic conditions, has also reported no MPOX cases. This study suggests that tropical and temperate climates are more conducive to MPOX relative to other climate types.

## RECOMMENDATIONS

Based on the study findings, we recommend that the World Health Organization, the Centers for Disease Control and Prevention, and national health agencies emphasize the importance of staying indoors in countries where MPOX is already prevalent, particularly on sunny days. This advice should apply to countries with precipitation values above 1500 mm and those with tropical and temperate climate types. Staying indoors will help reduce the spread of MPOX in conditions where contact is more likely and protect vulnerable populations, especially children. Practical advice on maintaining good physical and mental health while indoors should also be provided.

Additionally, tropical and temperate countries affected by MPOX should regularly monitor temperature changes, particularly in regions prone to temperature fluctuations. Implementing early warning strategies based on temperature trends in areas susceptible to MPOX would be beneficial.

Countries with precipitation values above 1500 mm should also enforce stricter health preventive measures and enhance health awareness to prevent the incidence or spread of MPOX.

There is a need to conduct localized research on the impact of climatic variations on MPOX prevalence at national and subcontinental levels, particularly in continents with extremely diverse climates.

We also emphasize the importance of a ring vaccination approach to curb the spread of MPOX, as recommended in a previous study [44]. Our findings highlight that those susceptible to the disease are often clustered within similar climatic conditions.

## LIMITATIONS

Linear regression assumes a linear relationship between the independent variables (AMSAT and precipitation) and the dependent variables (MPOX prevalence and mortality). This assumption may oversimplify the relationship if non-linear effects are present. However, the Prophet model accounts for seasonality and offers a robust alternative to manage this limitation.

There is also a potential for omitted variable bias, as the study focused solely on temperature and precipitation as predicting factors. Other variables may significantly influence MPOX prevalence. To address this bias, the study assessed the impact of adult literacy rate, net migration rate, and GDP per capita. Nonetheless, factors such as population density were not included in the analysis.

The findings may not be universally applicable to regions with highly diverse climates, such as the United States, Canada, Russia, and Argentina. In-depth, region-specific studies are needed in these countries to understand how climatic variations affect MPOX prevalence locally.

## CONCLUSION

The findings suggest that climate plays a significant role in MPOX dynamics, particularly in regions with less climatic diversity like South America and Europe. High temperatures, along with precipitation levels near and above 1500 mm, are particularly significant in the transmission of the MPOX virus. Additionally, tropical and temperate climates have been observed to be conducive to the spread of MPOX disease.

## Acknowledgement

No acknowledgement for this study.

## Author contributions statement

ISO made substantial contributions to the conceptualization of the study, acquisition, analysis, and interpretation of data, drafting of the first and final manuscript, and supervision of other authors; OJB made substantial contributions to the interpretation of data, drafting of the first and final manuscript. MAR made substantial contributions to the interpretation of data and drafting of the first and final manuscript. NA made substantial contributions to the interpretation of data and drafting of the first and final manuscript. OO made substantial contributions to the interpretation of data and drafting of the first and final manuscript. RCR made substantial contributions to the interpretation of data and drafting of the first and final manuscript. EJD made substantial contributions to the interpretation of data and drafting of the first and final manuscript. TO made substantial contributions to the interpretation of data and drafting of the first and final manuscript. IF made substantial contributions to the acquisition and analysis of data and drafting of the first and final manuscript. All authors read and approved the final manuscript. All authors agreed to be accountable for all aspects of the work.

## Data Availability Statement

The data that support the findings of this study are available from the corresponding author, ISO, upon reasonable request.

## References

1. WHO Director-General declares mpox outbreak a public health emergency of international concern. (2024). Accessed: Aug. 26: https://www.who.int/news/item/14-08-2024-who-director-general-declares-mpox-outbreak-a-public-health-emergency-of-international-concern

2. “What you need to know about monkeypox,” World Economic Forum. (2024). Accessed: Aug. 26: https://www.weforum.org/agenda/2024/08/monkeypox-virus-what-you-need-to-know-zoonosis-smallpox-public-health/.

3. Anil S, Joseph B, Thomas M, Sweety VK, Suresh N, and Waltimo T: Monkeypox: A Viral Zoonotic Disease of Rising Global Concern. 2024, 4:121–131. 10.1097/ID9.0000000000000124

4. Murhula Masirika L, Udahemuka JC, Ndishimye P, Sganzerla Martinez G, Kelvin P, Malyamungu Bubala N, Bilembo Kitwanda S, Kumbana Mweshi F, Mutimbwa Mambo L, B. Oude Munnink B, Bengehya Mbiribindi J: Epidemiology, clinical characteristics, and transmission patterns of a novel Mpox (Monkeypox) outbreak in eastern Democratic Republic of the Congo (DRC): an observational, cross-sectional cohort study. medRxiv. 2024, 2024:03. 10.1101/2024.03.05.24303395

5. Mathieu E, Spooner F, Dattani S, Ritchie H, Roser M: Mpox. Our World in Data. 2024.8.28

6. “2023 Outbreak in DRC,” Centers for Disease Control and Prevention. (2024). Accessed: Aug. 26: https://t.cdc.gov/001AR.

7. Mpox in DR Congo: The children who are suffering the most. (2024). Accessed: Aug. 26: https://www.bbc.com/news/articles/cdjwz77mmgmo.

8. Saied AA, Metwally AA, Choudhary OP. Monkeypox: an extra burden on global health. International journal of surgery. 2022 Aug 1;104:106745. 10.1016/j.ijsu.2022.106745

9. Fahrni ML, Sharma A, Choudhary OP. Monkeypox: prioritizing public health through early intervention and treatment. International Journal of Surgery. 2022 Aug 1;104:106774. 10.1016/j.ijsu.2022.106774

10. Nachega JB, Sam-Agudu NA, Ogoina D, Mbala-Kingebeni P, Ntoumi F, Nakouné E, Njouom R, Lewis RF, Gandhi M, Rosenthal PJ, Rawat A: The surge of mpox in Africa: a call for action. The Lancet Global Health. 2024, 12:e1086–e1088. 10.1016/S2214-109X(24)00187-6

11. Laurenson-Schafer H, Sklenovská N, Hoxha A, Kerr SM, Ndumbi P, Fitzner J, Almiron M, de Sousa LA, Briand S, Cenciarelli O, Colombe S: Description of the first global outbreak of mpox: an analysis of global surveillance data. The Lancet Global Health. 2023, 11(7):e1012–23. 10.1016/S2214-109X(23)00198-5

12. Musa-Booth TO, Medugu N, Adegboro B, Babazhitsu M: A review of the epidemiology, diagnosis, treatment, vaccines and economic impact of human monkeypox (Mpox) outbreaks. African Journal of Clinical and Experimental Microbiology. 2023, 24(1):1–8.

13. Pinto P, Costa MA, Gonçalves MF, Rodrigues AG, Lisboa C: Mpox person-to-person transmission—where have we got so far? a systematic review. Viruses. 2023, 15(5):1074. 10.3390/v15051074

14. Eser-Karlidag G, Chacon-Cruz E, Cag Y, Martinez-Orozco JA, Gudino-Solorio H, Cruz-Flores RA, Gonzalez-Rodriguez A, Martinez-Nieves D, Gomez-Zepeda M, Calderon-Suarez A, Çaşkurlu H: Features of Mpox infection: The analysis of the data submitted to the ID-IRI network. New Microbes and New Infections. 2023, 53:101154. 10.1016/j.nmni.2023.101154

15. Hazra A, Cherabie JN: Is Mpox a sexually transmitted infection? Why narrowing the scope of this disease may be harmful. Clinical Infectious Diseases. 2023, 76(8):1504–7. 10.1093/cid/ciac962

16. Mpox clade 1: what you need to know - UK Health Security Agency. (2024). Accessed: Aug. 25: https://ukhsa.blog.gov.uk/2024/08/23/mpox-clade-1-what-you-need-to-know/.

17. Choudhary OP, Chopra H, Shafaati M, Dhawan M, Metwally AA, Saied AA, Rabaan AA, Alhumaid S, Al Mutair A, Sarkar R. Reverse zoonosis and its relevance to the monkeypox outbreak 2022. New microbes and new infections. 2022 Nov;49. 10.1016/j.nmni.2022.101049

18. Fisman D: Seasonality of viral infections: mechanisms and unknowns. Clinical Microbiology and Infection. 2012, 18(10):946-954. 10.1111/j.1469-0691.2012.03968.x

19. Dai Q, Ma W, Huang H, Xu K, Qi X, Yu H, Deng F, Bao C, Huo X: The effect of ambient temperature on the activity of influenza and influenza like illness in Jiangsu Province, China. Science of the total environment. 2018, 645:684–691. 10.1016/j.scitotenv.2018.07.065

20. Azziz Baumgartner E, Dao CN, Nasreen S, Bhuiyan MU, Mah-E-Muneer S, Mamun AA, Sharker MY, Zaman RU, Cheng PY, Klimov AI, Widdowson MA: Seasonality, timing, and climate drivers of influenza activity worldwide. Journal of infectious diseases, vol. 206, no. 6. 2012, 206(6):838-46. 10.1093/infdis/jis467

21. Yamasaki L, Murayama H, Hashizume M: The impact of temperature on the transmissibility and virulence of COVID-19 in Tokyo, Japan. Sci Rep. 2021, 11(1):24477. 10.1038/s41598-021-04242-3

22. Shi P, Dong Y, Yan H, Li X, Zhao C, Liu W, He M, Tang S, Xi S: The impact of temperature and absolute humidity on the coronavirus disease 2019 (COVID-19) outbreak-evidence from China. MedRxiv. 2020, 2020–03. 10.1101/2020.03.22.20038919

23. Islam N, Bukhari Q, Jameel Y, Shabnam S, Erzurumluoglu AM, Siddique MA, Massaro JM, D’Agostino Sr RB: COVID-19 and climatic factors: A global analysis. Environmental Research. 2021, 193:110355. 10.1016/j.envres.2020.110355

24. World Economic Outlook (April 2024) - GDP per capita, current prices. (2024). Accessed: Aug. 26: https://www.imf.org/external/datamapper/NGDPDPC@WEO.

25. “Adult literacy rate,” Our World in Data. (2024). Accessed: Aug. 26: https://ourworldindata.org/grapher/literacy-rate-adults.

26. Choudhary OP, Fahrni ML, Saied AA, Chopra H. Ring vaccination for monkeypox containment: strategic implementation and challenges. International Journal of Surgery. 2022 Sep 1;105:106873. 10.1016/j.ijsu.2022.106873

27. Islam MA, Sangkham S, Tiwari A, Vadiati M, Hasan MN, Noor ST, Mumin J, Bhattacharya P, Sherchan SP: Association between global monkeypox cases and meteorological factors. International journal of environmental research and public health. 2022, 19(23):15638. 10.3390/ijerph192315638

28. World Health Organization. Joint ECDC-WHO Regional Office for Europe Mpox Surveillance Bulletin. (2024). Accessed: Aug. 26: https://monkeypoxreport.ecdc.europa.eu/.

29. WHO African Region Mpox Bulletin - 11 August 2024 - Democratic Republic of the Congo. (2024). Accessed: Aug. 19: https://reliefweb.int/report/democratic-republic-congo/who-african-region-mpox-bulletin-11-august-2024.

30. Home | Climate Change Knowledge Portal. (2024). Accessed: Aug. 26: https://climateknowledgeportal.worldbank.org/.

31. Koppen Climate Classification System. Accessed: Aug. 26: https://education.nationalgeographic.org/resource/koppen-climate-classification-system/.

32. World Bank Open Data. (2024). Accessed: Aug. 26: https://data.worldbank.org.

33. Net migration rate - The World Factbook. (2024). Accessed: Aug. 26: https://www.cia.gov/the-world-factbook/field/net-migration-rate/.

34. Peel MC, Finlayson BL, McMahon TA. Updated world map of the Köppen-Geiger climate classification. Hydrology and earth system sciences. 2007, 11(5):1633–44. 10.5194/hess-11-1633-2007

35. Ferreira GW, Reboita MS. A new look into the South America precipitation regimes: observation and forecast. Atmosphere. 2022, 13(6):873. 10.3390/atmos13060873

36. Bedson HS, Dumbell KR: The effect of temperature on the growth of pox viruses in the chick embryo. Epidemiology & Infection. 1961, 59(4):457–470. 10.1017/S0022172400039152

37. Bandyopadhyay S, Kanji S, Wang L: The impact of rainfall and temperature variation on diarrheal prevalence in Sub-Saharan Africa. Applied Geography. 2012, 33:63–72. 10.1016/j.apgeog.2011.07.017

38. Tagbo BN, Mwenda JM, Eke CB, Edelu BO, Chukwubuike C, Armah G, Seheri ML, Isiaka A, Namadi L, Okafor HU, Ozumba UC: Rotavirus diarrhoea hospitalizations among children under 5 years of age in Nigeria. Vaccine. 2018, 36(51):7759–7764. 10.1016/j.vaccine.2018.03.084

39. Dimitrova A, McElroy S, Levy M, Gershunov A, Benmarhnia T: Precipitation variability and risk of infectious disease in children under 5 years for 32 countries: a global analysis using Demographic and Health Survey data. Lancet Planetary Health. 2022, 6(2):e147–e155. 10.1016/S2542-5196(21)00325-9

40. Sharma A, Fahrni ML, Choudhary OP. Monkeypox outbreak: New zoonotic alert after the COVID-19 pandemic. International Journal of Surgery. 2022 Aug 1;104:106812. 10.1016/j.ijsu.2022.106812

41. Baker LB: Physiology of sweat gland function: The roles of sweating and sweat composition in human health. Temperature. 2019, 6(3):211–259. 10.1080/23328940.2019.1632145

42. Dhawan M, Choudhary OP. Emergence of monkeypox: risk assessment and containment measures. Travel Medicine and Infectious Disease. 2022 Sep;49:102392. 10.1016/j.tmaid.2022.102392

43. Park JH, Lee JW, Kim YC, Prausnitz MR: The effect of heat on skin permeability. Int J Pharm. 2008, 359(1-2):94–103. 10.1016/j.ijpharm.2008.03.032

44. Choudhary P, Shafaati M, Salah MA, Chopra H, Choudhary OP, Silva-Cajaleon K, Bonilla-Aldana DK, Rodriguez-Morales AJ. Zoonotic diseases in a changing climate scenario: Revisiting the interplay between environmental variables and infectious disease dynamics. Travel medicine and infectious disease. 2024 Feb 7:102694. 10.1016/j.tmaid.2024.102694

